# Plasma biomarkers and incident clinical stage progression in the specialized memory clinic: a longitudinal study

**DOI:** 10.64898/2026.10.19.26365317

**Authors:** Xuemei Zeng, Nicholas Heaton, Rebecca A. Deek, Michel N. Nafash, Marissa F. Farinas, Lamia T. Choity, Wasiu G. Balogun, Ruyu Shi, Margaret A. Bedison, Brian J. Lopresti, Cristy Matan, Neelesh K. Nadkarni, Alexandra Gogola, C. Elizabeth Shaaban, Howard J. Aizenstein, Dana L. Tudorascu, Tharick A. Pascoal, Victor L. Villemagne, Sarah B. Berman, Milos D. Ikonomovic, Robert A. Sweet, Julia K. Kofler, Beth E. Snitz, M. Ilyas Kamboh, Ann D. Cohen, Oscar L. Lopez, Thomas K. Karikari

**Affiliations:** Department of Psychiatry, School of Medicine, University of Pittsburgh, Pittsburgh, PA 15213, USA; Biofluid Biomarker Laboratory, Western Psychiatric Hospital, University of Pittsburgh Medical Center, Pittsburgh, PA 15213, USA; Alzheimer’s Disease Research Center, University of Pittsburgh, Pittsburgh, PA 15213, USA; Department of Biostatistics and Health Data Science, School of Public Health, University of Pittsburgh, Pittsburgh, PA 15213, USA; Department of Human Genetics, School of Public Health, University of Pittsburgh, Pittsburgh, PA, 15213, USA; Department of Radiology, School of Medicine, University of Pittsburgh, Pittsburgh, PA 15213, USA; Division of Geriatric Medicine, Department of Medicine, School of Medicine, University of Pittsburgh, Pittsburgh, PA 15213, USA; Department of Epidemiology, School of Public Health, University of Pittsburgh, Pittsburgh, PA, 15213, USA; Department of Health Promotion and Development, School of Nursing, University of Pittsburgh, Pittsburgh, PA, 15213, USA; Department of Neurology, School of Medicine, University of Pittsburgh, Pittsburgh, PA 15213, USA; Geriatric Research Education and Clinical Center, VA Pittsburgh HS, Pittsburgh, PA, USA; Department of Pathology, University of Pittsburgh, Pittsburgh, PA 15213, USA

**Keywords:** Alzheimer’s disease, blood biomarkers, amyloid beta, memory clinics, cognitive decline, clinical progression, Clinical Dementia Rating

## Abstract

**INTRODUCTION:** The utility of blood based Alzheimer’s disease (AD) biomarkers in predicting clinical trajectories remains uncertain.

**METHODS:** We assessed the diagnostic and prognostic performance of p-tau181, p-tau217, GFAP, and NfL in a large memory□clinic cohort (n=3,645) with comprehensive baseline clinical evaluation and annual follow□up for up to three decades.

**RESULTS:** P-tau217, p-tau181 and GFAP were associated with amyloid-beta (Aβ) pathology, while all biomarkers were elevated in AD and all-cause dementia. P-tau217 significantly outperformed others in detecting Aβ pathology and clinical AD but performed similarly to NfL and GFAP for all-cause dementia. Prognostically, p□tau217 demonstrated the strongest association with cognitive decline, followed by GFAP and NfL, and best predicted progression status at 5□ and 10□ year intervals. All biomarkers provided added prognostic value beyond demographic and genetic covariates.

**DISCUSSION:** These findings support plasma biomarkers for disease identification and risk stratification while underscoring the need for population□specific approaches.

## 1 Background

Alzheimer’s disease (AD) and AD-related dementias (ADRD) are the most prevalent forms of dementia, accounting for 60-80% of cases^1^. Pathologically, AD is hallmarked by the presence of amyloid beta (Aβ) plaques and neurofibrillary tangles (NFTs) consisting of highly phosphorylated tau (p-tau) protein^2^. Additionally, AD is characterized by significant synaptic and neuronal loss, particularly in brain regions critical for memory and cognitive function, such as hippocampus and cortex^3,4^.

Clinically, AD is characterized by progressive cognitive decline including memory loss, executive dysfunction, and impairments in language and visuospatial abilities^5^. However, the speed of progression varies significantly among individuals, influenced by factors such as age, lifestyle, educational attainment, and *APOE* ε4 genotype^6–8^. Given the profound impact of AD on patients and caregivers, and the recent approval of disease-modifying therapies such as donanemab and lecanemab^9,10^, identifying individuals at higher risk for rapid progression is critical for timely clinical intervention. Risk stratification is also essential for prioritizing access to these limited treatments, thereby maximizing their benefit-to-risk ratio.

In recent years, blood-based biomarkers have garnered significant attention as minimally invasive, scalable, and cost-effective tools for detecting AD pathophysiology^11–13^. Prominent emerging candidates include Aβ peptides (e.g., Aβ1-42/Aβ1-40 ratio)^14,15^, phosphorylated tau proteoforms (such as p-tau181 and p-tau217)^16–19^, the astrocytosis marker glial fibrillary acidic protein (GFAP)^20,21^, and the neuronal injury marker neurofilament light (NfL)^22,23^. Notably, the p-tau217/Aβ42 ratio was recently approved by the FDA as an in vitro diagnostic test for assessing Aβ pathology^24^. A growing body of evidence supports the association of these protein biomarkers with AD pathology^25–27^. Consequently, several of these biomarkers have been integrated into the revised diagnostic and staging criteria for AD by the Alzheimer’s Association working group^28,29^. Among these blood biomarkers, p-tau217 has superior diagnostic accuracy, with area under the curve (AUC) values typically exceeding 0.9 in studies distinguishing clinical or biological AD from cognitively unimpaired individuals^11,16,30,31^.

Despite their proven performance for biological AD, the utility of blood biomarkers in “real-world” clinical settings remains to be fully explored. A recent review of studies examining associations between blood biomarkers and cognitive performance reported that, although p tau biomarkers generally show robust correlations, results vary substantially across cohorts, cognitive assessment instruments, and specific cognitive domains^27^. This heterogeneity underscores the complex relationships between blood biomarkers and cognitive function and importantly, it highlights the need for continued investigation of biomarker–clinical associations in clinical settings, where patient populations are more heterogeneous and clinical decision making occurs, in order to establish the generalizability, interpretability, and clinical utility of blood biomarkers for routine implementation in Alzheimer’s disease care.

In this study, we investigated the association between baseline plasma biomarker levels and clinical stage progression over approximately three decades in a large memory clinic cohort across the spectrum of baseline normal cognition to dementia. By leveraging annual clinical assessments, we aimed to evaluate the potential utility of baseline plasma biomarkers as predictive tools in specialized memory clinics for estimating the time to progression to all-cause dementia and identifying high-risk individuals who may benefit from more definitive diagnostic testing.

## 2. METHODS

### 2.1 Study participants

This study included plasma samples from participants enrolled in the University of Pittsburgh Alzheimer’s Disease Research Center (PITT-ADRC) between 1993 and 2023. Inclusion criteria were aged 60 or older, or younger with self-reported symptoms of ADRD, > 7 years of education, adequate visual and auditory acuity to complete neuropsychological testing, and availability of reliable study partner. Exclusion criteria included history of major psychiatric disorders, recent electroconvulsive therapy, current substance related disorders, cancer within past 5 years (except skin and in situ prostate), or any significant medical condition that could affect neuropsychological testing. Blood was collected by venipuncture from participants at baseline visits using EDTA tubes and processed into plasma for biomarker measurements and buffy coat for *APOE* ε4 genotyping as described^32,33^. Additionally, participants underwent extensive baseline assessments, including neurologic, neuropsychological, functional, and psychiatric evaluations, laboratory assessment of vitamin B12 levels and thyroid function tests and brain magnetic resonance imaging (MRI).

Comprehensive sociodemographic information, including sex, self-reported race, and years of education, was also collected. Participants were followed for annual clinical assessments and, in some cases, additional blood draws. Cognitive adjudications were performed at consensus meetings that included review of all clinical information, blood work, and brain MRI. A detailed description of the participant recruitment and various clinical assessments was previously published^34^. The PITT-ADRC study was approved by the University of Pittsburgh Institutional Review Board (STUDY 19110245). All participants provided written consent. A flowchart depicting sample selection is presented in Figure S1.

Clinical progression was assessed using the Clinical Dementia Rating Global Score (CDR-GS), a widely used tool in clinical and community settings for diagnosing and staging AD^35,36^. Participants were stratified according to their baseline CDR-GS as cognitively normal (CN; CDR-GS=0), mild cognitive impairment (MCI; CDR-GS=0.5), and dementia (CDR-GS>=1). Due to the limited number of participants of other races, only participants self-identified as non-Hispanic White (NHW) or Black/African-American (B/AA) were included in the statistical analysis.

Positron emission tomography (PET) imaging of Aβ pathology was acquired from a subset of participants using Pittsburgh Compound B (¹¹C PiB) as a radiotracer, as previously described^37–40^. The acquired scans were processed using CapAIBL^41^ to derive MR-independent Centilloid (CL)-scaled indices of Aβ burden. Participants were dichotomized into A+ or A- using a positivity threshold of 20 CL.

### 2.2 Plasma biomarker measurement

Plasma biomarker measurements were conducted in the Biomarker Biofluid Laboratory, Department of Psychiatry, University of Pittsburgh, using commercially established Single Molecule Array (Simoa) assay kits on HD-X instruments (Quanterix, Billerica, MA, USA). Plasma samples were thawed and centrifuged at 4000xg for 10 minutes to remove particulates before instrument analysis. The p-tau181 and p-tau217 levels were measured using the p-tau181 V2 Advantage (#103714) and ALZpath p-Tau 217 V2 (#104371), respectively. NfL and GFAP were measured using Neuro 2-Plex B (#103520). Three quality control (QC) samples were run at the beginning and end of each run to monitor assay precision. The mean within-run coefficient of variation (CV) and between-run CV were 4.2% and 6.9% for p-tau181, 8.1% and 6.9% for p-tau217, 6.5% and 8.9% for NfL, and 7.3% and 9.4% for GFAP.

### 2.3 Statistical analysis

Data analysis was conducted using MATLAB (R2024a) and R statistical software v4.2.1 (R Foundation for Statistical Computing, Vienna, Austria). Demographic characteristics were summarized as medians with interquartile ranges (IQRs) for continuous variables and as counts for categorical variables. Group differences were assessed using the Kruskal-Wallis (continuous) or Chi-square test (categorical). Participants with no follow-up CDR-GS or a baseline CDR-GS of 3 were excluded from the longitudinal analyses. Benjamini–Hochberg procedure was used for false discovery rate adjustment^42^. Covariate adjustment refers to the addition of age, sex, race, education, and *APOE* ε4 carrier status to the models.

Biomarker values were log_2_-transformed to reduce skewness prior to statistical analysis. An analysis of covariance (ANCOVA) was used to examine baseline cross-sectional associations between biomarkers and the CDR-GS or clinical diagnosis, using Cohen’s f-statistic^43^ as the effect size measure. For two group comparisons, effect sizes were quantified using Cohen’s d^43^. The area under the receiver operating characteristic (ROC) curve (AUC) was used to quantify the performance of biomarkers in differentiating between two groups. AUC confidence intervals (CI) were estimated via bootstrap (1,000 replicates). DeLong tests were used to compare AUC^44^. Youden index was used to determine the optimal classification threshold.

Time-to-incident clinical stage progression was defined as the period between blood draw and the first visit with a CDR-GS higher than the baseline. Participants with no CDR-GS increase were censored on their last visit. Cox proportional hazards regression models were used to examine the association between biomarkers and time to incident progression. Biomarkers were analyzed as either continuous variables or categorized into quartiles. In the continuous models, biomarker levels were log transformed, and hazard ratios (HRs) were expressed per one standard deviation (SD) increase in the log transformed values. To assess potential effect modification by covariates, interaction terms between biomarkers and covariates of interest were included in the models. Covariate stratified hazard ratios were derived from these interaction models. For quartile-based analyses, cutoffs were defined based on the distribution of biomarker levels in the full cohort. Kaplan-Meier methods were used to visualize survival curves by quartile and estimate median survival times. Incidence rates of clinical progression were compared across quartiles using Poisson regression models with a log link, with person□years of follow□up included as an offset. P values were derived from Wald tests of regression coefficients comparing each quartile with the reference group (Q1).

Participants were classified as progressors over 5- or 10-year periods if their CDR-GS increased during the specified timeframe. Non-progressors showed no increase in CDR-GS within the period and the first visit thereafter. Logistic regression models were used to evaluate the effectiveness of biomarkers in discriminating progressors from non-progressors, with effect sizes quantified using Cohen’s d and performance evaluated using ROC analysis.

The associations of plasma biomarkers with longitudinal trajectories of CDR sum of box (CDR-SB) were examined using linear mixed effects models with random intercepts and random slopes for each participant. The models included fixed effects for time from blood draw, dichotomized biomarker group (defined as above or below median of the full cohort), and their interaction, along with covariates.

## 3 RESULTS

### 3.1 Participant characteristics

This study included 3,645 baseline plasma samples from participants enrolled at PITT-ADRC. Samples were selected based on the availability of complete measurements for four core AD plasma biomarkers (p tau181, p tau217, GFAP, and NfL) and a baseline CDR GS obtained within six months of the blood draw (Figure S1). Participants self reported a race other than NHW or B/AA were excluded because of small sample sizes (n = 23). Among the 3,645 included participants, the median age was 73.4 years (IQR, 12.9); 59.6% were female, 90.8% were NHW, 57.6% had at least some college education, and 48.0% were *APOE* ε4 carriers.

Participant characteristics by CDR-GS are shown in Table 1. Older age, female sex, NHW race, being *APOE* ε4 carriers, and lower education were associated with higher CDR-GS (lower cognitive function). CDR-GS generally matched clinical diagnoses. Most participants with a CDR GS of 0 (93.8%) were clinically normal. Among participants with a CDR GS ≥ 1, the majority were clinically diagnosed with dementia, including 85.9%–86.4% with AD dementia and 11.1%–13.6% with non AD dementia across the three CDR GS groups. Participants with a CDR-GS of 0.5 represented a more heterogeneous group, spanning all four diagnostic categories. Because of the long historical timeframe of evaluations (1993-current), some diagnostic practices changed over time, likely accounting for some variability (e.g., MCI was not a diagnostic entity until early 2000s). Among the 3,645 participants, 250 underwent antemortem *in vivo* assessment of Aβ pathology using ¹¹C□PiB PET, with 116 and 134 participants classified as negative (A-) and positive (A+) according to CapAIBL CL cutoff of 20 (Table S1)^41^. A total of 2,457 had follow-up CDR assessments, with a median follow-up duration of 3.0 years (IQR 1.9-5.6 years; maximum: 26.1 years).

**Table 1.**
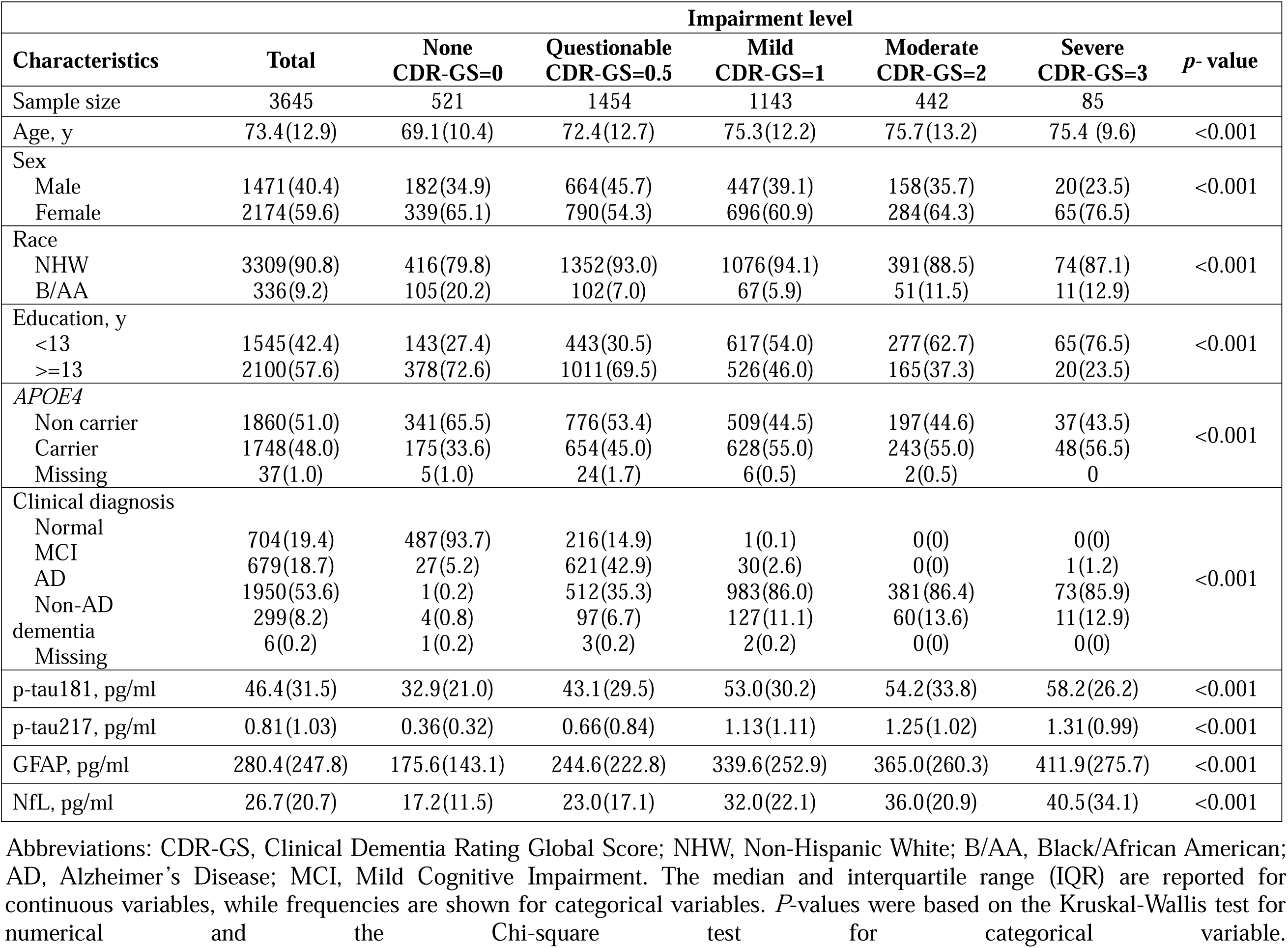
Characteristics of the study cohort.

### 3.2 Diagnostic performance for identifying brain amyloid-beta pathology

We first evaluated the diagnostic performance of plasma biomarkers in identifying brain amyloidosis assessed by ¹¹C□PiB PET. As showed in Figure 1A, plasma p□tau217 showed the largest group level difference between A+ and A-participants, followed by GFAP, p□tau181, and NfL. The corresponding effect sizes were 2.28 (95% CI 2.01–2.56; p□<□0.001), 1.13 (95% CI 0.86–1.41; p□<□0.001), 0.84 (95% CI 0.57–1.11; p□<□0.001), and 0.26 (95% CI −0.01–0.53; p□=□0.060), respectively, after adjustment for the covariates (age, sex, race, *APOE* ε4 carriership, and education).

**Figure 1.**
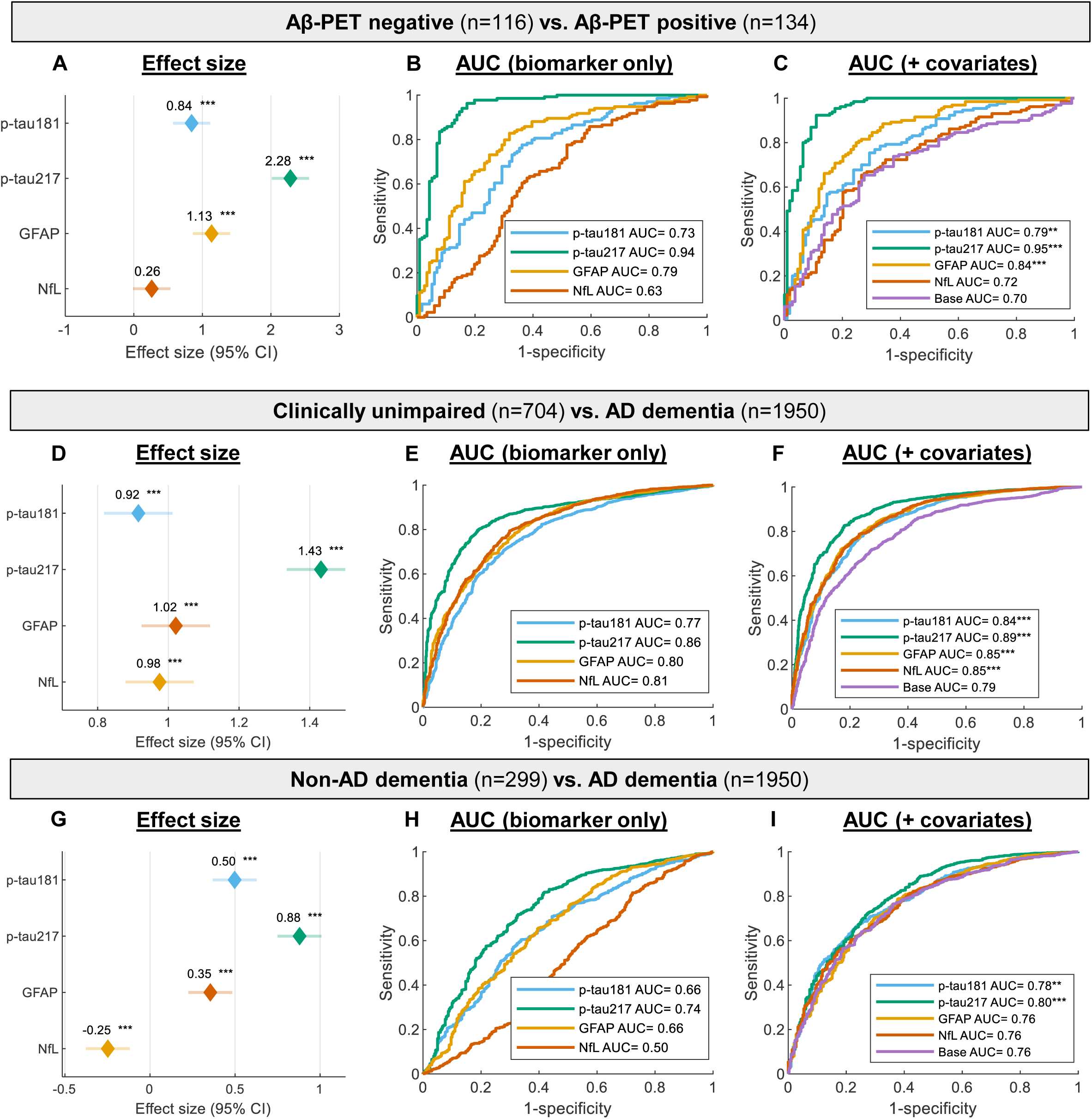
Diagnostic performance of plasma biomarkers for discriminating Aβ-PET positivity (A) or clinical AD (B-C). Panels A, D, and G show Cohen’s d effect sizes with 95% confidence intervals, adjusted for covariates; biomarker levels were z normalized, and p values were false discovery rate (FDR)–adjusted using the Benjamini–Hochberg method. Panels B, E, and H present ROCs from logistic regression models using biomarkers alone, whereas panels C, F, and I include covariates. Asterisks in panels C, F, and I indicate DeLong test comparisons versus the covariate-only (base) model. *, **, and *** denote p < 0.05, 0.01, and 0.001, respectively.

Consistently, p-tau217 had the highest AUC in classifying Aβ-PET status. In biomarker□alone models (Figure 1B), p□tau217 achieved an AUC of 0.94 (95% CI 0.91–0.97), which was significantly higher than that of GFAP (AUC 0.79, 95% CI 0.73–0.85), p□tau181 (AUC 0.73, 95% CI 0.66–0.79), and NfL (AUC 0.63, 95% CI 0.55–0.69), with all DeLong tests p < 0.001. Inclusion of covariates (Figure 1C) slightly improved the discriminative performance over p□tau217 alone (AUC 0.95, 95% CI 0.92–0.97; DeLong p=0.049), which was significantly better than the covariate□only model (AUC 0.70, 95% CI 0.64-0.76; DeLong p < 0.001). Similarly, inclusion of p□tau181 or GFAP significantly improved the covariate□only model, yielding covariate□adjusted AUCs of 0.79 (95% CI 0.72–0.84; DeLong p = 0.003) and 0.84 (95% CI 0.78–0.88; DeLong p < 0.001), respectively. In contrast, inclusion of NfL did not significantly improve model performance.

Figures S2A depicts the boxplot distribution of plasma biomarkers by Aβ-PET status, which showed the unadjusted fold change of 1.51, 3.13, 1.70 and 1.10, for p-tau181, p-tau217, GFAP and NfL, respectively, with rank sum p values < 0.001 for all four biomarkers. Additional performance metrics for plasma biomarker–based discrimination between A+ and A-individuals are provided in Tables□S2-3. Consistent with its superior discriminative performance, p□tau217 achieved high negative predictive value (NPV; 95.0%) and positive predictive value (PPV; 87%) as a standalone biomarker.

### 3.3 Diagnostic performance for identifying clinical AD dementia

All four plasma biomarkers significantly differentiated clinically unimpaired (CU) individuals from those with AD dementia (Figure□1D–F). P□tau217 demonstrated the largest covariate□adjusted effect size (1.43; 95% CI 1.33–1.53), followed by GFAP (1.02; 95% CI 0.92–1.12), NfL (0.98; 95% CI 0.88–1.07), and p□tau181 (0.92; 95% CI 0.82–1.01). In terms of classification performance, p□tau217 achieved an AUC of 0.86 (95% CI 0.84–0.87) in biomarker□only models, which increased to 0.89 (95% CI 0.88–0.90) after covariate adjustment. In both models, p□tau217 significantly outperformed p□tau181, GFAP, and NfL (all DeLong tests, p < 0.001). GFAP and NfL showed comparable discriminative performance, with biomarker□only AUCs of 0.80 (95% CI 0.79–0.82) and 0.81 (95% CI 0.79–0.82), respectively, both significantly higher than that of p□tau181 (AUC = 0.77; 95% CI 0.75–0.79), with DeLong p values of 0.004 and 0.002, respectively. After covariate adjustment, AUCs for GFAP and NfL increased to 0.85 (95% CI 0.83–0.87), which were no longer significantly different from that of p□tau181 (0.84; 95% CI 0.82–0.86). The addition of any of the four plasma biomarkers to the covariate□only model (AUC = 0.79; 95% CI 0.77–0.80) significantly improved discriminative performance. Across biomarkers, discrimination between CU and AD was characterized by high PPVs but relatively lower NPVs, with PPVs ranging from 0.87–0.92 and NPVs from 0.47–0.59 in biomarker□only models, increasing to 0.90–0.93 and 0.56–0.62, respectively, after inclusion of covariates (Tables□S2–3).

When differentiating non□AD dementia from AD dementia (Figure 1G–I), p□tau217, p□tau181, and GFAP showed positive covariate adjusted effect sizes, indicating higher concentrations in individuals with AD dementia. Effect sizes ranged from 0.35 to 0.88, with GFAP showing the smallest and p□tau217 the largest effect. In contrast, NfL exhibited a negative covariate adjusted effect size (−0.25, 95% CI [-0.37, −0.12]), indicating higher levels in individuals with non□AD dementia. In terms of classification performance, p-tau217 again demonstrated the best performance, with AUC of 0.74 (95% CI 0.71–0.77), followed by GFAP and p-tau181, both with AUCs of 0.66. NfL showed no significance in the biomarker-only model. Inclusion of p-tau181 (DeLong p=0.001) or p-tau217 (DeLong p<0.001), but not GFAP and NfL improved over the performance of the covariate-only model. However, the magnitude of increase was relatively small, increased from covariate-only AUC of 0.76 (95% CI 0.72–0.78) to 0.78 (95% CI 0.75–0.80) with the inclusion of p-tau181 and 0.80 (95% CI 0.77–0.82) for p-tau217. Similar to the discrimination between CU and AD, all four biomarkers demonstrated high PPVs but low NPVs when differentiating non AD dementia from AD, with overall performance lower than those observed for CU versus AD.

Figure S2B show the boxplot distributions of biomarker levels by clinical diagnosis. P-tau217 exhibited the highest magnitude of covariate-adjusted difference across groups, with effect sizes of 0.45 (95% CI 0.28–0.88). This was followed by NfL (0.34; 95% CI 0.22–0.68), GFAP (0.32; 95% CI 0.20–0.63), and p-tau181 (0.30; 95% CI 0.18–0.56). All four biomarkers exhibited a significant stepwise increase from CU to MCI and to AD dementia.

### 3.4 Diagnostic performance for identifying all-cause dementia

P□tau217 exhibited the largest effect size (1.23; 95% CI 1.13–1.34) when comparing participants with all□cause dementia (defined as CDR-GS ≥ 1) to cognitively normal individuals (CDR-GS = 0), followed by NfL, GFAP, and p□tau181 (Figure 2A). However, the magnitude of differences among p□tau217, NfL, and GFAP was substantially reduced (effect size ranging from 1.12 to 1.23) compared with those observed for Aβ pathology and clinical AD dementia described above. In terms of discriminative performance, both p tau217 and NfL achieved an AUC of 0.83 (95% CI 0.81–0.85), slightly higher than that of GFAP (AUC = 0.81; 95% CI 0.78–0.83; DeLong p values of 0.022 for p-tau217 and 0.076 for NfL; Figure 2B). All three biomarkers showed significantly better performance than p tau181 (AUC = 0.76; 95% CI 0.74–0.78; all DeLong p values < 0.001). Inclusion of covariates increased AUCs to a range of 0.82–0.86 (Figure 2C). All four biomarkers significantly improved discrimination between cognitive unimpaired and all-cause dementia when included to the covariate□only model (AUC = 0.76; 95% CI 0.74–0.79). As observed for clinical AD dementia, all biomarkers demonstrated high PPVs but relatively low NPVs (Tables□S2–3).

**Figure 2.**
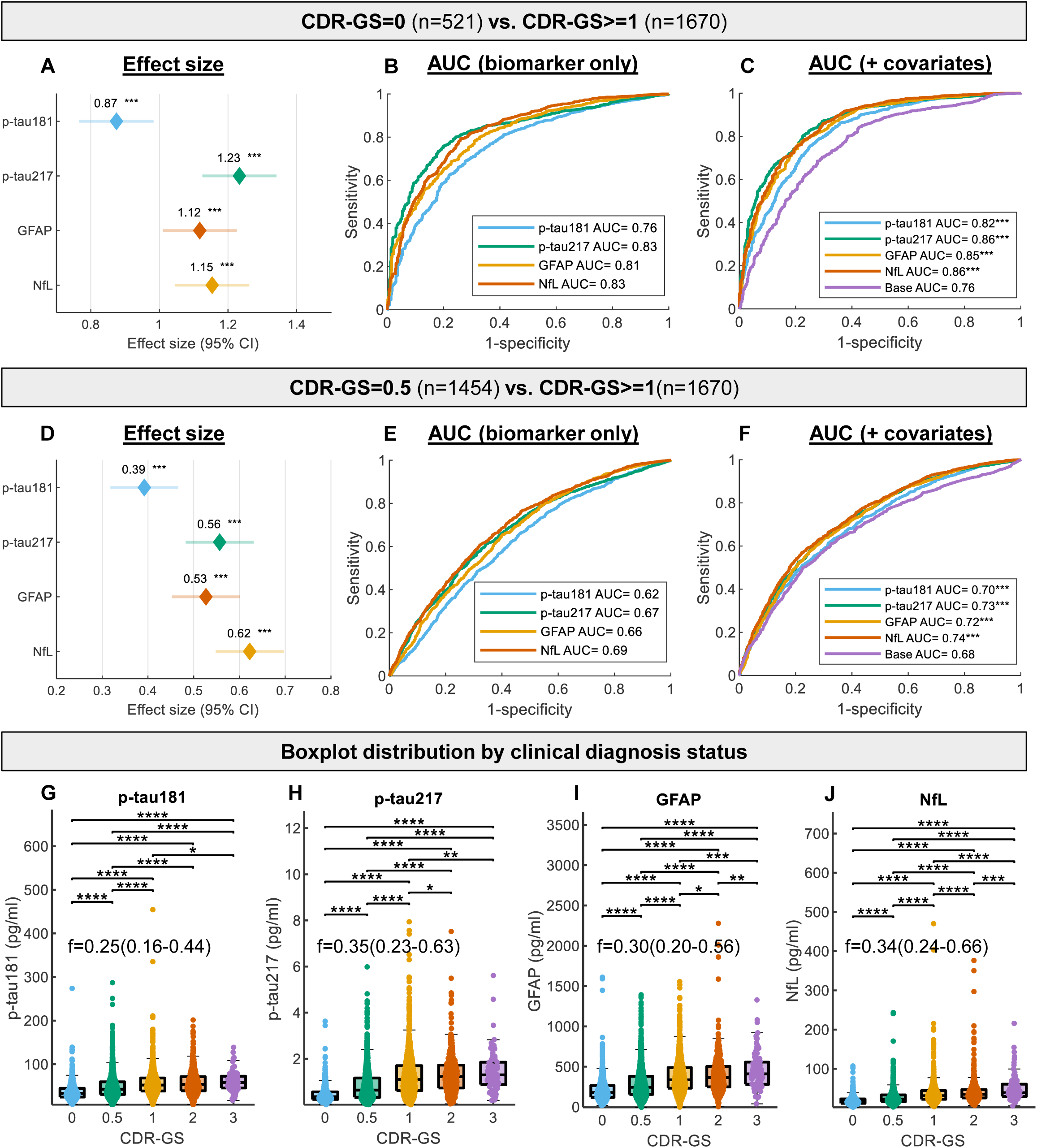
Diagnostic performance of plasma biomarkers for all-cause dementia. Panels A–C distinguish baseline CDR□GS = 0 vs ≥1; panels D–F compare CDR□GS = 0.5 vs ≥1. Panels A and D show covariate□adjusted Cohen’s d (95% CI); biomarker levels were z□normalized. Panels B and E present ROC curves from biomarker only models, whereas panels C and F include covariates (asterisks denote DeLong comparisons versus the base model). Panels G–J show boxplots by CDR□GS (median, IQR, and whiskers to non□outliers). One extreme value each for p□tau217 and NfL is omitted for visualization only. P values are from Wilcoxon rank□sum tests with Benjamini–Hochberg correction. Cohen’s f summarizes effect size across groups. *, **, and *** denote P < 0.05, 0.01, 0.001.

Effect sizes comparing participants with CDR-GS-defined MCI (CDR-GS = 0.5) to those with all cause dementia were attenuated, ranging from 0.39 to 0.62, with the largest effect observed for NfL, followed by p□tau217, GFAP, and p□tau181 (Figure 2D). In terms of discriminative performance, NfL also demonstrated the highest AUC (0.69; 95% CI 0.67–0.71), although this was not significantly higher than that of p□tau217 (AUC = 0.67; 95% CI 0.65–0.69; DeLong p value = 0.09; Figure 2E). Inclusion of covariates modestly improved performance, increasing AUCs to a range of 0.70–0.74 (Figure 2F). PPVs ranged from 0.62–0.67 and NPVs from 0.59–0.66 for biomarker-only models (Table S3). Corresponding ranges increased to 0.69–0.74 for PPVs and 0.61–0.64 for NPVs after inclusion of covariates.

Figure 2G–J show the boxplot distributions of plasma biomarker levels across CDR-GS categories. P-tau217 and NfL exhibited the strongest covariate-adjusted associations, with effect sizes of 0.35 (95% CI 0.23–0.63) and 0.34 (95% CI 0.24–0.66), respectively. These were followed by GFAP (0.30; 95% CI 0.20–0.56), and p-tau181 (0.25; 95% CI 0.16–0.44). All four plasma biomarkers exhibited significant stepwise increases from CDR□GS 0 to 0.5 and from 0.5 to 1 (all p < 0.001). Beyond a CDR□GS of 1, NfL demonstrated a more pronounced stepwise increase compared with the other three biomarkers.

### 3.5 Prognostic utility for clinical stage progression

We next assessed the performance of plasma biomarkers in predicting progression across clinical stages, defined as an increase in CDR□GS. Among all participants with follow□up visits (participant characteristics summarized in Table S4), and treating biomarker levels as continuous variables, all four biomarkers were significantly associated with time-to-clinical progression (Table 2). HRs per SD were highest for p□tau217 (1.80; 95% CI 1.69–1.92), followed by GFAP (1.70; 95% CI 1.58–1.83), NfL (1.67; 95% CI 1.56–1.79), and p□tau181 (1.37; 95% CI, 1.29–1.46), after adjustment for baseline covariates. All associations were statistically significant (p<0.001).

**Table 2.** Association of plasma biomarkers with time to incident clinical stage progression.

|  | Full cohort<br>N=2457 |  | Normal (CDR-GS=0)<br>N=378 |  | MCI (CDR-GS=0.5)<br>N=1096 |  | Dementia (CDR-GS$\geq 1$)<br>N=983 |  |
| --- | --- | --- | --- | --- | --- | --- | --- | --- |
|  | HR | p value | HR | p value | HR | p value | HR | p value |
| **Biomarker-only models** |  |  |  |  |  |  |  |  |
| p-tau181 | 1.44(1.36-1.53) | <0.001 | 1.42(1.17-1.73) | <0.001 | 1.34(1.23-1.47) | <0.001 | 1.20(1.09-1.31) | <0.001 |
| p-tau217 | 1.91(1.80-2.03) | <0.001 | 2.50(1.95-3.20) | <0.001 | 1.88(1.72-2.05) | <0.001 | 1.29(1.17-1.42) | <0.001 |
| GFAP | 1.83(1.72-1.96) | <0.001 | 1.96(1.55-2.46) | <0.001 | 1.75(1.58-1.93) | <0.001 | 1.28(1.15-1.42) | <0.001 |
| NfL | 1.74(1.64-1.85) | <0.001 | 1.68(1.33-2.12) | <0.001 | 1.67(1.53-1.83) | <0.001 | 1.30(1.17-1.43) | <0.001 |
| **Adjusted for covariates** |  |  |  |  |  |  |  |  |
| p-tau181 | 1.37(1.29-1.46) | <0.001 | 1.26(1.02-1.56) | 0.032 | 1.27(1.15-1.39) | <0.001 | 1.22(1.11-1.34) | <0.001 |
| p-tau217 | 1.80(1.69-1.92) | <0.001 | 2.29(1.73-3.02) | <0.001 | 1.75(1.59-1.93) | <0.001 | 1.29(1.17-1.43) | <0.001 |
| GFAP | 1.70(1.58-1.83) | <0.001 | 1.59(1.24-2.06) | <0.001 | 1.51(1.35-1.70) | <0.001 | 1.30(1.16-1.46) | <0.001 |
| NfL | 1.67(1.56-1.79) | <0.001 | 1.38(1.04-1.83) | 0.024 | 1.54(1.38-1.71) | <0.001 | 1.37(1.23-1.52) | <0.001 |

Abbreviation: CDR-GS, Clinical Dementia Rating global score; HR, hazard ratio. Hazard ratios were estimated using Cox proportional hazards models and standardized to the standard deviation of log<sub>10</sub>-transformed biomarker values. Unadjusted models were based solely on the biomarker, and adjusted models included covariates for age, sex, race, *APOE4* carrier status, and education level ( $\geq 13$ years vs. $< 13$ years).

When stratified by baseline impairment status, p□tau217 retained the strongest association with clinical progression among participants who were cognitively unimpaired (CDR GS=0) or had MCI (CDR□GS=0.5), but not among those with dementia (CDR-GS>=1), with covariate adjusted HRs per SD of 2.29 (95% CI 1.73–3.02), 1.75 (95% CI 1.59–1.93), and 1.29 (95% CI 1.17–1.43), respectively (Table 2). Associations for both p tau217 and GFAP were significant across all cognitive stages, being strongest for baseline cognitively normal individuals. In contrast, p tau181 and NfL showed only nominal significance among cognitively unimpaired individuals after covariate adjustment (unadjusted p < 0.024).

Similarly, stratification by p□tau217 quartiles demonstrated the greatest separation in incident clinical progression rates, median survival time (MST), and HRs (Figure 3). Participants in the highest quartile (Q4) had an incident rate (IR) of 29.75 per 100 person□years and an MST of 2.17 years, whereas those in the lowest quartile (Q1) had a markedly lower IR of 5.87 per 100 person□years and a substantially longer MST of 14.4 years. The covariate□adjusted HR for Q4 relative to Q1 was 4.62 (95% CI 3.79–5.64). NfL exhibited the second□strongest separation, followed by GFAP and then p tau181 (Figure 3B-F). Notably, when comparing Q2 with Q1, NfL showed superior predictive performance relative to the other biomarkers, with a covariate□adjusted hazard ratio of 2.31 (95% CI 1.91–2.79).

**Figure 3.**
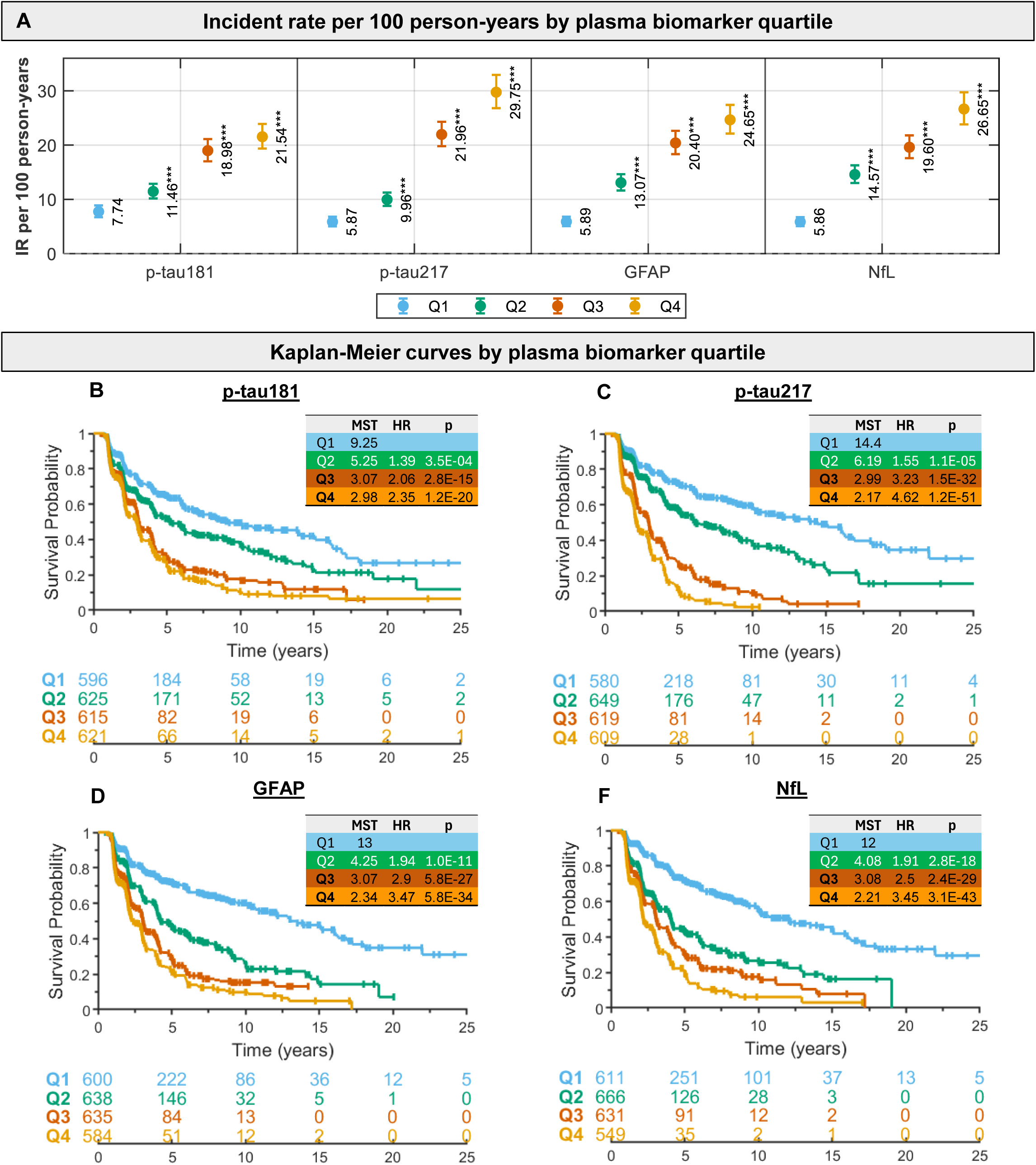
Prognostic utility of plasma biomarkers for clinical stage progression. Panel A shows forest plots of incident rates (IRs) per 100 person years across biomarker quartiles. Asterisks indicate differences relative to the reference quartile (Q1), based on Poisson regression models with a log link and person years of follow up included as an offset. *, **, and *** denote p < 0.05, 0.01, and 0.001, respectively. Panels B–F display Kaplan–Meier curves illustrating associations between plasma biomarkers and time to clinical progression. Hazard ratios (HRs) and p values were estimated using covariate□adjusted Cox proportional hazards models. Survival probability reflects the likelihood of remaining cognitively stable (i.e., no increase in CDR□GS). MST denotes median stability time.

### 3.6 Differentiating clinical stage progressors from non-progressors

We conducted ROC analyses to evaluate the utility of baseline plasma biomarkers for predicting clinical progression over a 5□year period (Figure□4). Table□S5 summarizes the characteristics of participants with available 5□year progression status, as defined in the Methods. Older age, female sex, NHW race, lower education attainment, and *APOE* ε4 carriership were associated with higher odds of progression.

P□tau217 again showed the strongest discrimination between progressors and non□progressors, with an effect size of 1.10 (95% CI 0.97–1.23) and a biomarker□only AUC of 0.82 (95% CI 0.80–0.84) when all participants with available 5□year progression data were included (Figure 4A–B). NfL and GFAP demonstrated comparable performance, with effect sizes of 0.96 (95% CI 0.82–1.09) and 0.92 (95% CI 0.79–1.06), respectively, and AUCs of approximately 0.78. P□tau181 showed the lowest performance, with an effect size of 0.69 (95% CI 0.56–0.83) and an AUC of 0.73 (95% CI 0.70–0.76). Inclusion of covariates improved discrimination across all biomarkers, increasing AUCs to a range of 0.80 to 0.85, all of which were significantly higher than the covariate□only model (Figure 4C).

**Figure 4.**
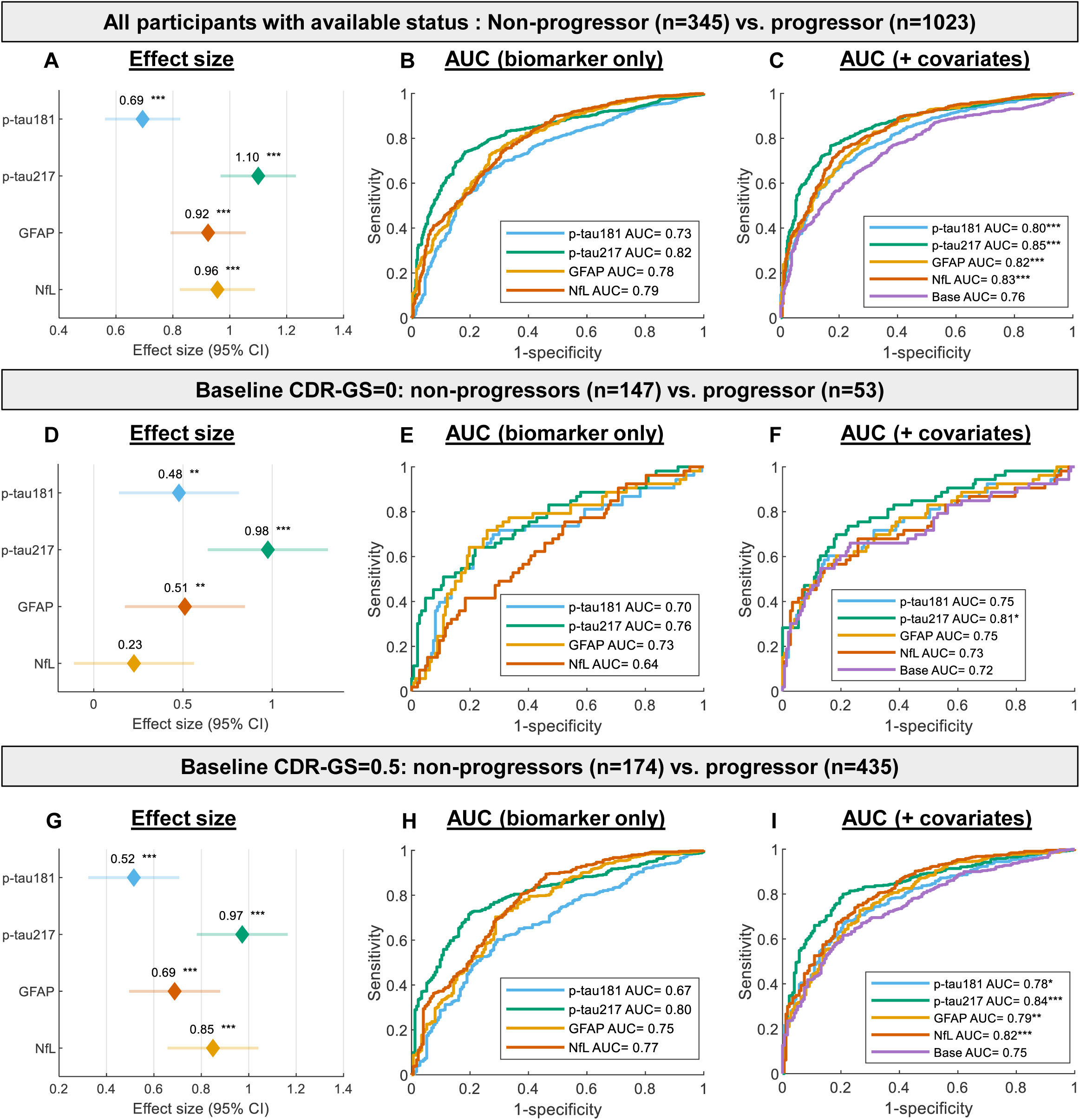
Performance in differentiating 5□year progression status. Panels A–C show biomarker performance in distinguishing individuals who progressed within 5 years (progressors) from those who remained stable for at least 5 years (non□progressors). Panels D–I present performance stratified by baseline CDR□GS. Panels A, D, and G show covariate□adjusted Cohen’s d effect sizes with 95% confidence intervals; biomarker levels were z□normalized, and p values were FDR□adjusted using the Benjamini–Hochberg method. Panels B, E, and H show ROC curves from logistic regression models using biomarkers alone, whereas panels C, F, and I show models additionally adjusted for covariates (asterisks denote DeLong test comparisons versus the covariate□only model). *, **, and *** denote p < 0.05, 0.01, and 0.001, respectively.

When examining performance among participants who were cognitively unimpaired (CDR□GS□=□0; Figure□4D–F) or mildly impaired at baseline (CDR□GS□=□0.5; Figure□4G–I), p□tau217 remained the strongest predictor in both groups. Overall, all four biomarkers demonstrated better discriminative performance among participants with baseline MCI compared with those who were cognitively normal. Notably, NfL showed no significant association with 5□year progression status among cognitively unimpaired individuals after covariate adjustment. In this group, p tau217 was the only biomarker whose addition to the covariate only model significantly improved discriminative performance (DeLong p□=□0.029).

Similar findings were observed when evaluating baseline plasma biomarkers for differentiating 10□year progressors from non progressors (Supplementary Figure S3 and Table□S5). Overall, biomarkers demonstrated better performance for predicting 10□year progression than 5□year progression, with biomarker only AUCs ranging from 0.72 to 0.87 (Figure S3). NfL showed performance comparable to p tau217 when all participants with available 10□year progression status were included and among individuals with MCI at baseline. Similar to 5-year progression status, p-tau217 was the only biomarker whose addition to covariate-model showed significant improvement (DeLong p=0.015).

All four biomarkers showed high PPVs but relatively low NPVs for distinguishing progressor status at both 5□and 10□year intervals (Tables S2–3), with PPVs ranging from 0.88–0.92 and 0.97–0.98, and NPVs ranging from 0.42–0.51 and 0.17–0.29, respectively.

### 3.7 Association with longitudinal CDR-SB trajectories

We next evaluated the performance of baseline plasma biomarkers in predicting longitudinal trajectories of CDR□SB, which provides a more granular measure of cognitive and functional severity assessment. Higher baseline levels of all four biomarkers (dichotomized by the population median) were associated with faster increases in CDR□SB, indicating more rapid cognitive and functional decline (Figure 5A). P□tau217 showed the largest difference in slope (Δslope) between participants with high versus low levels, with Δslope of 1.05 (±0.05), followed by GFAP (0.83±0.05), NfL (0.75±0.05), and p□tau181 (0.71±0.05). All associations were significant, with covariate-adjusted p values < 0.001.

**Figure 5.**
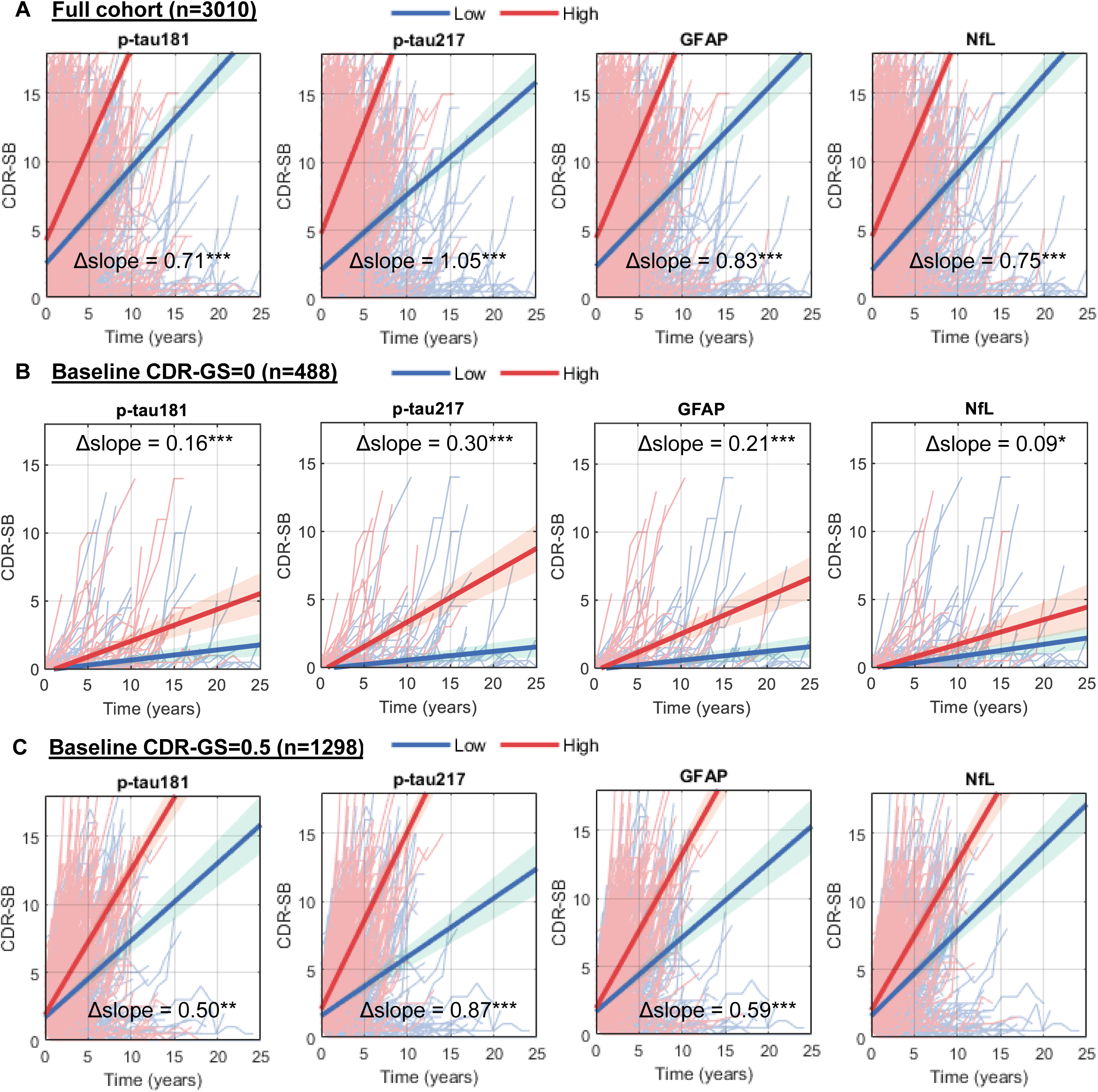
Performance in predicting longitudinal trajectory of Clinical Dementia Rating Sum of Box (CDR-SB). Spaghetti plots depict individual CDR SB trajectories over time, stratified by baseline plasma biomarker levels dichotomized at the population median, for the full cohort (A), participants with baseline CDR□GS□=□0 (B), and participants with baseline CDR□GS□=□0.5 (C). Slope differences were estimated using linear mixed effects models with CDR SB as the outcome and biomarker group as the predictor, including participant level random intercepts and adjustment for covariates. *, **, and *** denote p□<□0.05, 0.01, and 0.001, respectively. Low and high refer to below median and above median, respectively.

**Figure 6.**
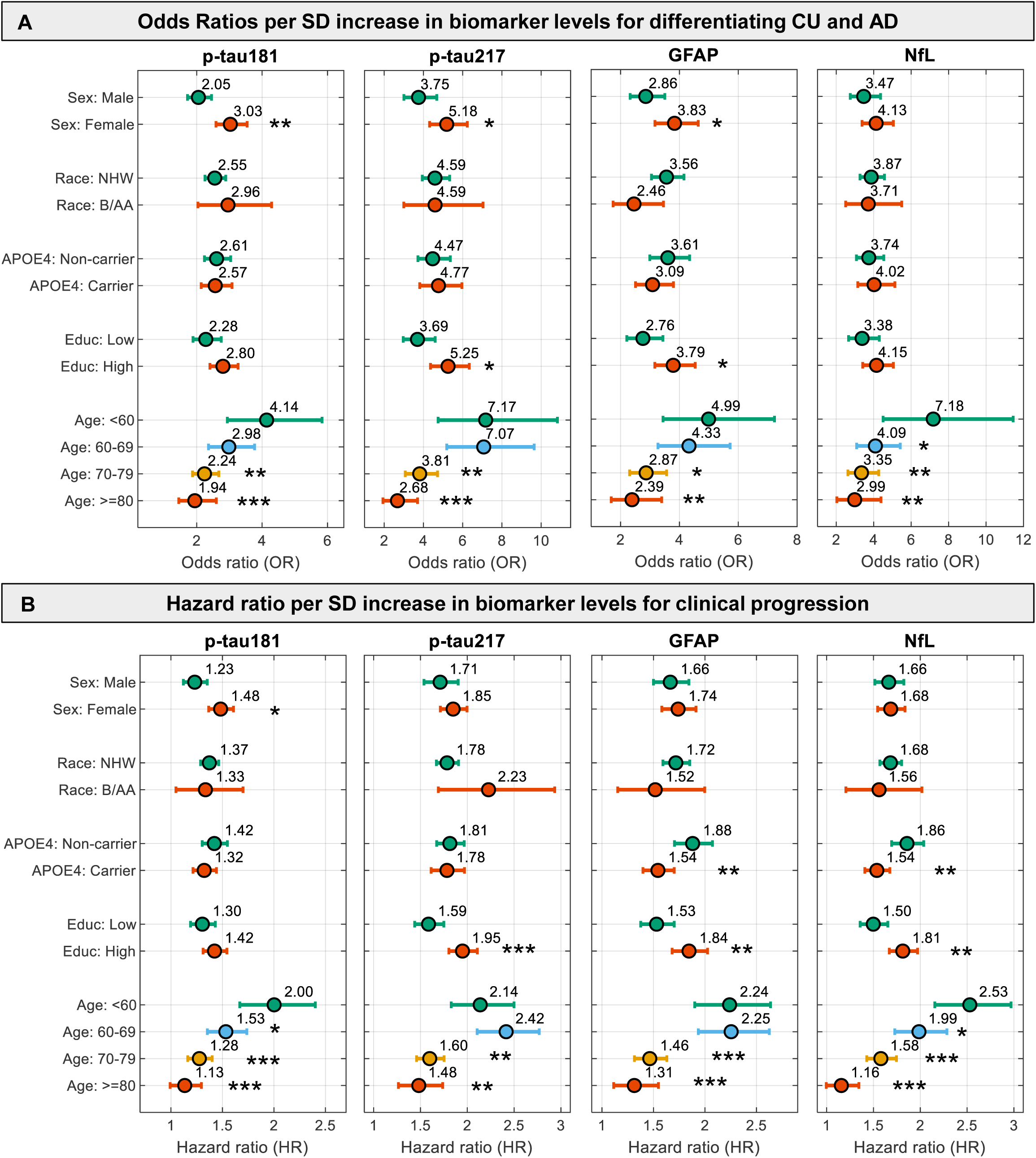
Impact of covariates in biomarker performance. Panel A presents forest plots of odds ratios (ORs) per standard deviation (SD) increase in plasma biomarkers for differentiating clinically unimpaired (CU) individuals from those with AD dementia. ORs and 95% CIs were derived from multivariable logistic regression models including log□□transformed, z□normalized plasma biomarkers, the covariate of interest, and the corresponding biomarker × covariate interaction term, with adjustment for all remaining covariates. Panel B illustrates forest plots of hazard ratios (HRs) for clinical progression by plasma biomarker levels. HRs and 95% CIs were derived from Cox proportional hazards models including log□□transformed, z normalized plasma biomarkers, the covariate of interest, and the corresponding biomarker × covariate interaction term, with adjustment for all remaining covariates. Asterisks in both panels denote statistically significant interaction effects relative to the reference categories (age <60 years; sex, male; race, non Hispanic White [NHW]; *APOE4* non carrier; education <13 years). *, **, and *** denote p□<□0.05, 0.01, and 0.001, respectively.

When stratified by baseline cognitive impairment status, p□tau217 continued to exhibit the largest Δslope among both cognitively unimpaired individuals (CDR-GS=0; Figure□5B) and those with CDR-GS-defined MCI at baseline (Figure□5C), with Δslopes of 0.30 (±0.04) and 0.87 (±0.06), respectively. Consistent with its weaker association with longitudinal clinical stage progression among individuals without baseline impairment, NfL showed only nominal significance for predicting longitudinal CDR□SB trajectories in cognitively unimpaired participants, with a small Δslope of 0.09□±□0.04 (p□=□0.019).

## 4 DISCUSSION

Several blood biomarkers, including the four evaluated in this study (p□tau181, p□tau217, GFAP, and NfL), have demonstrated strong associations with AD. The four biomarkers were selected because they have consistently demonstrated their utility in diagnosing AD pathology. Notably, p-tau217 and p-tau181 have already received FDA approval for this purpose. However, further evaluation in large and particularly longitudinal cohorts is critically needed to establish their clinical utility in routine practice. A key strength of this study is the use of a large academic memory clinic cohort (PITT□ADRC) with extensive longitudinal follow□up spanning up to three decades and comprehensive clinical characterization. This unique design provided a robust real world framework to rigorously assess the performance of these four core AD plasma biomarkers for diagnosing Aβ pathology, clinical AD dementia, and all□cause dementia as defined by CDR□GS. In addition, we evaluated their prognostic utility for predicting clinical stage progression and cognitive trajectories, as well as the influence of key demographic and genetic factors on biomarker performance.

Overall, consistent with prior studies, p□tau217 demonstrated the strongest performance across most diagnostic and prognostic metrics evaluated^45–48^. Its highest discriminative accuracy was observed for Aβ-PET positivity, where it differentiated A+ from A-individuals with an AUC of 0.94, achieving 96% sensitivity, 83% specificity, and 90% accuracy with a single Youden index–derived cutpoint. These findings align with previously reported performance^46^, and approach the recommended performance threshold of ≥90% sensitivity and specificity for confirmatory testing^49,50^. Although associations remained strong for clinical outcomes, discriminative performance was progressively lower for clinical AD dementia and all□cause dementia. This pattern is consistent with the notion that p tau217 is most strongly associated with AD□specific pathology^28^, and its diagnostic accuracy may be reduced in more heterogeneous clinical settings due to potential misclassification of clinical AD and the presence of non□AD etiologies within all□cause dementia.

When Aβ-PET positivity was used as the outcome, p□tau217, p□tau181, and GFAP showed significant associations, whereas NfL did not. NfL is widely recognized as a non□specific marker of axonal injury that is elevated in AD as well as other neurodegenerative conditions^51,52^. While NfL levels have consistently been associated with future cognitive decline^53^, evidence for its cross sectional association with Aβ pathology has been mixed, with some studies reporting significant associations^47^, and others finding no relationship^54,55^. In our study, the lack of association between NfL and Aβ-PET positivity likely reflects the inclusion of individuals with non□AD dementia. After excluding these participants, a significant positive association between NfL and Aβ-PET positivity emerged (data not shown). This suggests that NfL’s relationship with amyloid burden is sensitive to cohort composition, with mixed etiologies in heterogeneous clinical populations potentially obscuring this association.

When stratified by clinical AD diagnosis, all four plasma biomarkers showed a stepwise increase across the AD continuum and were able to distinguish clinically unimpaired individuals from those with AD dementia, with each adding value beyond covariates. In contrast, when comparing non-AD dementia from AD-dementia, NfL, unlike the other biomarkers, did not differentiate non□AD dementia from AD dementia and was higher in non□AD dementia after covariate adjustment. This pattern is consistent with its role as a non□specific marker of neurodegeneration, with levels that may be elevated across diverse non□AD etiologies.

All four biomarkers also showed a stepwise increase with the severity of cognitive impairment as assessed by CDR□GS and demonstrated significant discriminative ability for distinguishing cognitively normal individuals from those with dementia, as well as individuals with MCI from those with dementia. However, when all cause cognitive impairment severity was used as the outcome, the relative advantage of p□tau217 was notably reduced. In contrast to analyses using Aβ-PET positivity and clinical AD diagnosis as outcomes, where p□tau217 consistently outperformed the other biomarkers, p□181, NfL, and GFAP showed comparable covariate adjusted effect sizes across CDR□GS comparisons. Notably, NfL exhibited slightly stronger associations than p□tau217 when differentiating MCI from dementia, although this difference was not statistically significant. These findings are consistent with the notion that plasma p tau217 is an early biomarker of AD□specific pathology^26,56^, and that diagnostic performance for clinical cognitive impairment may be moderated by the temporal lag between the emergence of pathology and the onset of cognitive symptoms^28,57^.

All four biomarkers demonstrated prognostic utility across multiple outcomes, including CDR□GS–defined clinical stage progression, 5□ and 10□year progression status, and longitudinal CDR□SB trajectories, consistent with prior studies^58–62^. Similar to their performance for all cause dementia, although p tau217 maintained the strongest overall performance, the differences among p tau181, GFAP, and NfL were relatively modest when analyses were conducted across all participants (excluding those with baseline CDR□GS of 3).

Notably, greater differences emerged when analyses were stratified by baseline cognitive status. NfL showed limited predictive robustness for clinical stage progression, 5□year progression, and CDR□SB trajectories among participants who were cognitively normal at baseline. In contrast, p□tau217 demonstrated the strongest prognostic performance in this group, with the highest HRs per SD increase observed among cognitively normal individuals, and progressively attenuated effects in those with MCI and dementia. These findings align with the existing literature, which indicates that p□tau217 levels become abnormal early in AD, while NfL typically rises later in both AD and other neurodegenerative diseases^56,63–65^. Together, these observations suggest that p□tau217, but not NfL, may be better suited for detecting early AD pathology and identifying individuals at high risk for progression prior to the onset of clinical symptoms.

It is worth noting that although we observed AUC values for discriminating progressors and non progressors that were comparable to those reported by others in research cohorts^61,62,66^, our results differed in predictive value profiles. Specifically, whereas prior studies have reported higher NPVs than PPVs, we observed high PPVs and relatively low NPVs. This discrepancy underscores the importance of considering population heterogeneity in the clinical implementation of plasma AD biomarkers. In contrast to research cohorts, which often include more preclinical participants, our cohort comprised individuals attending a memory clinic, many of whom were symptomatic or had subjective cognitive complaints. Consistent with this, a majority of participants progressed within 10 years, which likely influenced the observed predictive value patterns. Nevertheless, our findings suggest that in a memory clinic setting, plasma biomarkers may be better suited for identifying individuals at higher risk of cognitive decline rather than serving as rule out tests for low risk.

There are some limitations in our study. First, the memory clinic setting may limit the generalizability of the findings to broader or more diverse populations. Although age was the only exclusion criterion for study enrollment, our cohort includes a higher proportion of participants with baseline cognitive impairments, with nearly 50% exhibiting mild to severe dementia. Second, our analysis was limited to NHW and B/AA due to the limited number of participants from other racial and ethnic groups. Third, all our biomarker measurements were conducted in a single batch with reagents from controlled lots. Therefore, our results did not account for the additional variation introduced by the need for measurement at the time of collection.

In summary, leveraging a large memory clinic cohort with extensive long term follow□up, our study provides a comprehensive real world evaluation of plasma AD biomarkers and demonstrates their utility for AD diagnosis as well as their association with incident clinical progression. These findings support the integration of plasma biomarkers into clinical practice for risk stratification, early intervention planning, and personalized patient management.

## Data Availability

All data produced in the present study are available upon reasonable request to the authors

## ACKNOWLEDGMENTS

The authors are thankful to the Pitt-ADRC participants and study coordinators whose dedication and commitment were fundamental to the successful implementation of this study, MIK and members of the MIK Laboratory for the storage of ADRC samples over the past 30 years.

## CONSENT STATEMENT

All the studies were approved by the Institutional Review Board at the University of Pittsburgh. All subjects provided written informed consent.

## RESEARCH IN CONTEXT

- **Systematic review:** Using PubMed and Google Scholar as search engines, the authors focus on articles investigating the role of biomarkers in cognitive decline and clinical progression. Prior studies have often focused on single biomarkers or have been conducted in volunteer-based cohorts and have limited follow-up. Our study extends this work by examining multiple biomarkers in a longitudinal memory clinic cohort with data spanning up to three decades.
- **Interpretation:** All four biomarkers—p tau181, p tau217, GFAP, and NfL—were elevated in both Alzheimer’s disease and all-cause dementia, with performance influenced by common demographic factors. P tau217 showed the strongest association with worsening cognitive decline, followed by GFAP and NfL. Its advantage was more pronounced for identifying biological and clinical Alzheimer’s disease but less evident all-cause dementia.
- **Future directions:** Future studies involving more diverse racial and ethnic groups, along with long-term follow-up in community settings and real-time biomarker measurement are needed to improve generalizability.

## HIGHLIGHTS

- We assessed four key blood AD biomarkers in a large memory clinic cohort with long term follow-up.
- P□tau217 outperformed p□tau181, GFAP, and NfL in detecting dementia and predicting progression.
- P□tau217 showed greatest advantage for Aβ pathology, less for clinical AD and all-cause dementia.
- Biomarker elevations tracked worse trajectories; p□tau217 best separated progression risk.
- Biomarker performance was influenced by demographic factors, particularly age and education.

