## Supplementary Information for "Plasma biomarkers and incident clinical stage progression in the specialized memory clinic: a longitudinal study"

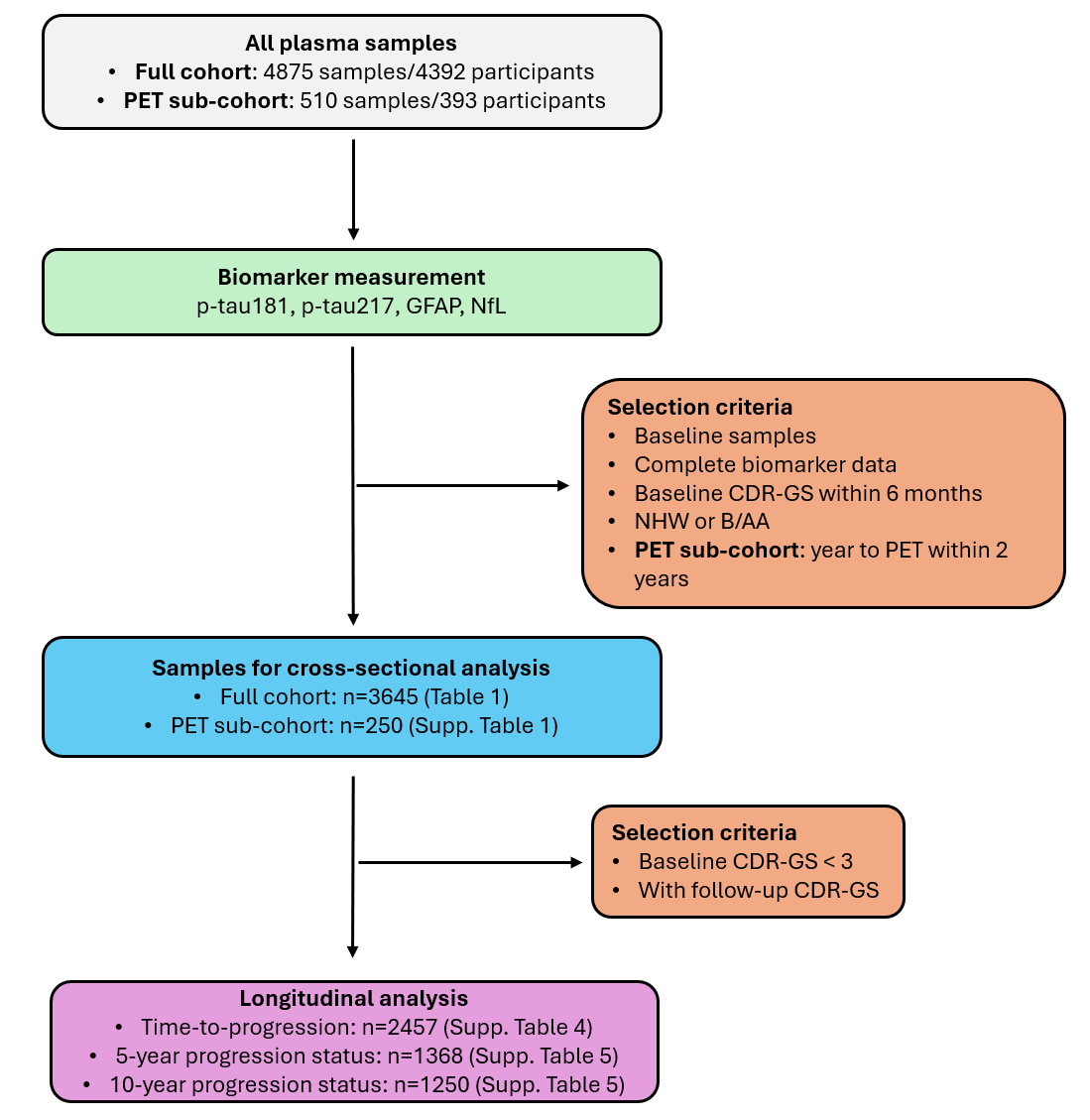

**Figure S1: Flow chart illustrating the sample selection process.** The diagram outlines the criteria used to select plasma samples from PITT-ADRC for this study. CDR-GS: Clinical Dementia Rating Global Score; NHW, non-Hispanic White; B/AA, Black/African American. MCI, mild cognitive impairment; PET, positron emission tomography; Supp., Supplementary Information. Characteristics for participants included in the analysis are summarized in the tables indicated in parentheses.

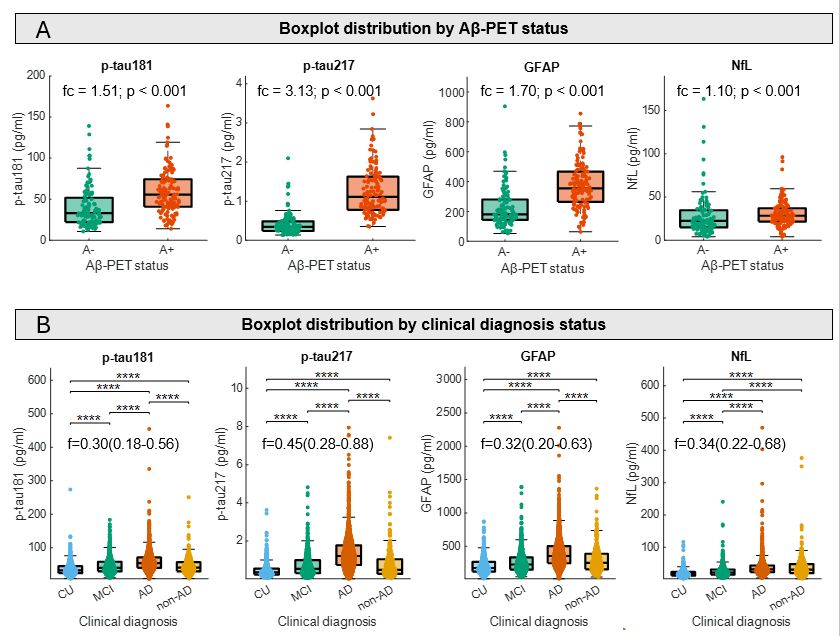

**Figure S2. Boxplot distribution of plasma biomarkers by Aβ-PET (A) or clinical diagnosis (B).** Center lines indicate medians, box edges represent the 25th and 75th percentiles, and whiskers extend to the most extreme non‑outlier values. One extreme value each for p‑tau181, p‑tau217, and GFAP was not shown in the boxplots but was included in the statistical analyses. Wilcoxon rank‑sum tests with Benjamini–Hochberg correction were used for significance assessment. *, **, and *** denote p < 0.05, 0.01, and 0.001, respectively.

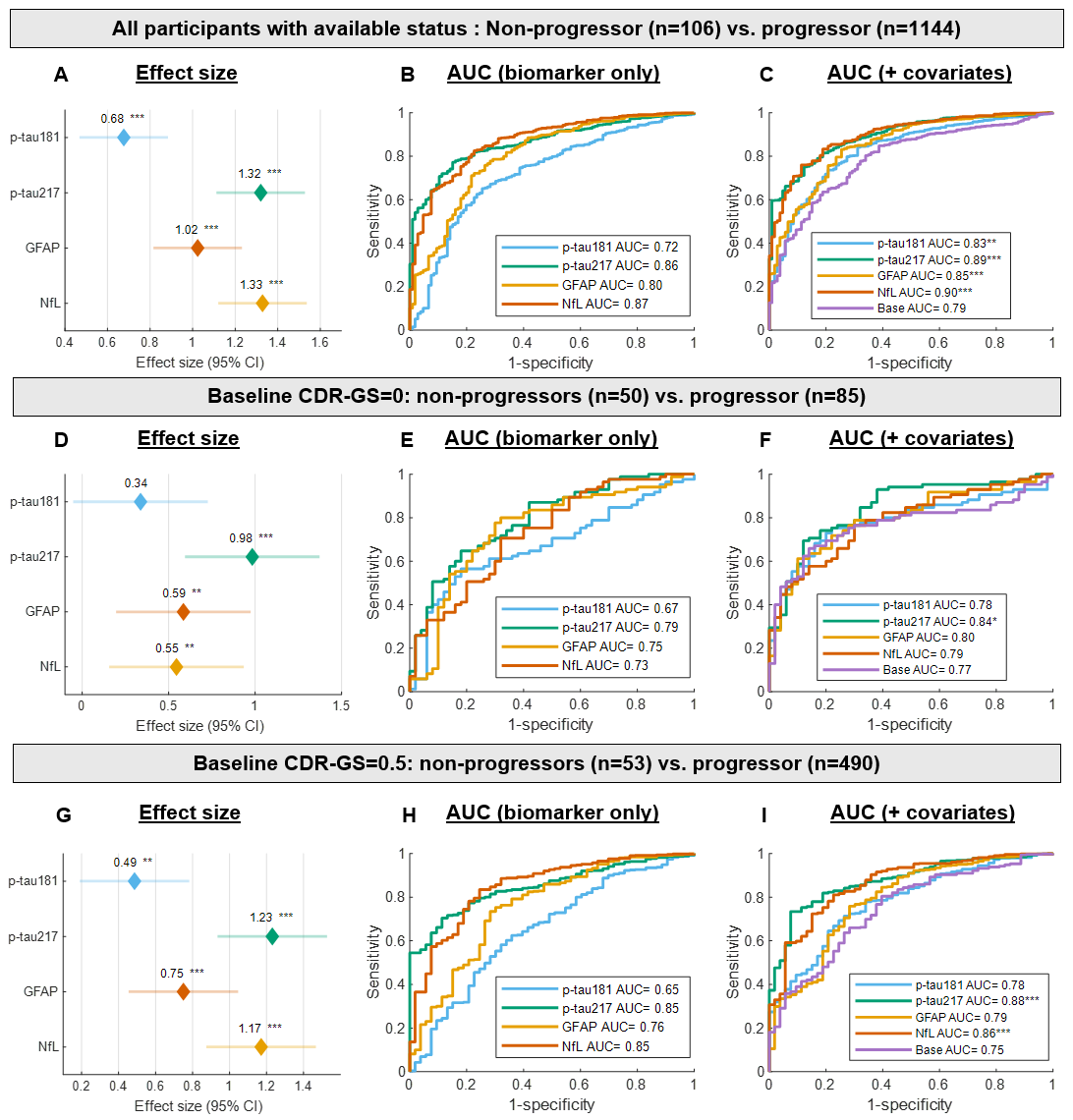

**Figure S3: Performance in differentiating 10‑year progression status.** Panels A–C show biomarker performance in distinguishing individuals who progressed within 10 years (progressors) from those who remained stable for at least 10 years (non‑progressors) among all participants with available progression status. Panels D–F and Panels G–I present corresponding comparisons among participants with baseline CDR‑GS = 0 (Panels D–F) and CDR‑GS = 0.5 (Panels G–I), respectively. Panels A, D, and G display Cohen’s d effect size estimates with 95% confidence intervals, adjusted for covariates; biomarker levels were z‑normalized, and p values were false discovery rate (FDR)–adjusted using the Benjamini–Hochberg procedure. Panels B, E, and H show area under the curve (AUC) values from logistic regression models using biomarker levels alone, whereas Panels C, F, and I show AUCs from models additionally adjusted for covariates (asterisks denote DeLong test comparisons with the covariate‑only model). Covariates included age, sex, race, APOE4 carriership, and education (<13 vs ≥13 years). *, **, and *** denote p < 0.05, 0.01, and 0.001, respectively.

**Table S1.** **Characteristics of the PET sub-cohort.**

| **Characteristics** | **Total** | **A-** | **A+** | **p value** |
| --- | --- | --- | --- | --- |
| Sample size | 250 | 116 | 134 |  |
| Age, y | 68.5(12.8) | 65.5(12.2) | 70.3(11.9) | 8.01E-04 |
| Sex |  |  |  | 0.729 |
| Male | 143(57.2) | 65(56.0) | 78(58.2) |  |
| Female | 107(42.8) | 51 (44.0) | 56(41.8) |  |
| Race |  |  |  | 0.905 |
| NHW | 241(96.4) | 112(96.6) | 129(96.3) |  |
| B/AA | 9(3.6) | 4(3.4) | 5(3.7) |  |
| Education, y |  |  |  | 0.237 |
| <13 | 42(16.8) | 16(13.8) | 26(19.4) |  |
| >=13 | 208(83.2) | 100(86.2) | 108(80.6) |  |
| *APOE4* |  |  |  | 1.04E-05 |
| Non carrier | 141(56.4) | 81(69.8) | 60(44.8) |  |
| Carrier | 98(39.2) | 28(24.1) | 70(52.2) |  |
| Missing | 11(4.4) | 7(6.0) | 4(3.0) |  |
| Clinical diagnosis |  |  |  | 1.16E-11 |
| Normal | 63(25.2) | 44(37.9) | 19(14.2) |  |
| MCI | 85(34.0) | 45(38.8) | 40(30.0) |  |
| AD | 77(30.8) | 10(8.6) | 67(50.0) |  |
| Non-AD dementia | 24(9.6) | 16(13.8) | 8(6.0) |  |
| Missing | 1(0.4) | 1(0.9) | 0(0) |  |
| CDR-GS |  |  |  | 6.53E-04 |
| 0 | 39(15.6) | 24(20.7) | 15(11.2) |  |
| 0.5 | 155(62.0) | 78(67.2) | 77(57.5) |  |
| 1 | 45(18.0) | 9(7.8) | 36(26.7) |  |
| 2 | 11(4.4) | 5(4.3) | 6(4.5) |  |
| p-tau181, pg/ml | 44.7(34.6) | 33.0(29.7) | 55.5(33.5) | 2.21E-10 |
| p-tau217, pg/ml | 0.67(0.88) | 0.34(0.25) | 1.11(0.85) | 1.55E-33 |
| GFAP, pg/ml | 279.1(223.6) | 179.5(136.7) | 354.3(203.5) | 1.36E-15 |
| NfL, pg/ml | 25.5(17.8) | 22.4(19.8) | 28.3(15.4) | 5.56E-04 |

Abbreviations: PET, positron emission tomography; CDR-GS, Clinical Dementia Rating Global Score; NHW, Non-Hispanic White; B/AA, Black/African American; AD, Alzheimer’s Disease; MCI, Mild Cognitive Impairment. The median and interquartile range (IQR) are reported for continuous variables, while frequencies are shown for categorical variables. P-values were based on the Wilcoxon rank sum test for numerical and the Chi-square test for categorical variable.

**Table S2. Plasma biomarker performance for Aβ‑PET positivity, clinical diagnosis, all‑cause dementia, and 5‑ or 10‑year progression (biomarker‑only models).**

|  | **AUC (95% CI)** | **Threshold** | **OR** | **NPV** | **PPV** | **Sens.** | **Spec.** | **Accu.** |
| --- | --- | --- | --- | --- | --- | --- | --- | --- |
| **Aβ-PET positive vs. Aβ-PET negative** | | | | | | | | |
| p-tau181 | 0.73(0.66-0.79) | 39.85 | 6.11 | 0.71 | 0.71 | 0.78 | 0.64 | 0.71 |
| p-tau217 | 0.94(0.91-0.97) | 0.55 | 123.84 | 0.95 | 0.87 | 0.96 | 0.83 | 0.90 |
| GFAP | 0.79(0.73-0.85) | 232.26 | 10.30 | 0.78 | 0.75 | 0.83 | 0.68 | 0.76 |
| NfL | 0.63(0.55-0.69) | 18.83 | 4.12 | 0.71 | 0.63 | 0.86 | 0.41 | 0.65 |
| **Clinically unimpaired vs. AD dementia (Biomarker-only models)** | | | | | | | | |
| p-tau181 | 0.77(0.75-0.79) | 43.18 | 6.27 | 0.47 | 0.88 | 0.71 | 0.72 | 0.71 |
| p-tau217 | 0.86(0.84-0.88) | 0.66 | 16.97 | 0.59 | 0.92 | 0.80 | 0.81 | 0.80 |
| GFAP | 0.80(0.78-0.82) | 231.14 | 7.88 | 0.53 | 0.87 | 0.79 | 0.68 | 0.76 |
| NfL | 0.81(0.79-0.83) | 21.46 | 9.06 | 0.55 | 0.88 | 0.79 | 0.70 | 0.77 |
| **Non-AD dementia vs. AD dementia** | | | | | | | | |
| p-tau181 | 0.66(0.62-0.69) | 48.83 | 2.99 | 0.20 | 0.92 | 0.60 | 0.66 | 0.61 |
| p-tau217 | 0.74(0.71-0.77) | 0.63 | 6.25 | 0.33 | 0.93 | 0.82 | 0.58 | 0.79 |
| GFAP | 0.66(0.63-0.70) | 283.58 | 2.90 | 0.21 | 0.92 | 0.66 | 0.60 | 0.65 |
| NfL | 0.50(0.46-0.55) | 20.64 | 1.70 | 0.19 | 0.88 | 0.82 | 0.28 | 0.74 |
| **CDR-GS=0 vs. CDR-GS>=1** | | | | | | | | |
| p-tau181 | 0.76(0.74-0.78) | 43.35 | 5.77 | 0.42 | 0.89 | 0.69 | 0.72 | 0.70 |
| p-tau217 | 0.83(0.81-0.85) | 0.67 | 12.61 | 0.51 | 0.92 | 0.76 | 0.80 | 0.77 |
| GFAP | 0.81(0.79-0.83) | 231.14 | 8.04 | 0.50 | 0.89 | 0.78 | 0.69 | 0.76 |
| NfL | 0.83(0.81-0.85) | 22.44 | 10.80 | 0.53 | 0.91 | 0.79 | 0.74 | 0.78 |
| **CDR-GS=0.5 vs. CDR-GS>=1** | | | | | | | | |
| p-tau181 | 0.62(0.60-0.64) | 43.28 | 2.29 | 0.59 | 0.62 | 0.69 | 0.51 | 0.60 |
| p-tau217 | 0.67(0.65-0.69) | 0.96 | 3.00 | 0.60 | 0.67 | 0.61 | 0.65 | 0.63 |
| GFAP | 0.66(0.65-0.68) | 284.17 | 2.82 | 0.60 | 0.65 | 0.66 | 0.60 | 0.63 |
| NfL | 0.69(0.67-0.71) | 23.79 | 3.68 | 0.66 | 0.65 | 0.77 | 0.53 | 0.66 |
| **Progressors vs. non-progressors (5-year period)** | | | | | | | | |
| p-tau181 | 0.73(0.70-0.76) | 43.17 | 5.33 | 0.42 | 0.88 | 0.66 | 0.73 | 0.68 |
| p-tau217 | 0.82(0.80-0.84) | 0.65 | 12.74 | 0.51 | 0.92 | 0.74 | 0.82 | 0.76 |
| GFAP | 0.78(0.76-0.81) | 237.39 | 7.27 | 0.48 | 0.89 | 0.73 | 0.72 | 0.73 |
| NfL | 0.79(0.76-0.81) | 21.40 | 6.44 | 0.48 | 0.88 | 0.75 | 0.69 | 0.73 |
| **Progressors vs. non-progressors (10-year period)** | | | | | | | | |
| p-tau181 | 0.72(0.67-0.77) | 42.90 | 5.46 | 0.17 | 0.97 | 0.65 | 0.75 | 0.66 |
| p-tau217 | 0.86(0.83-0.89) | 0.53 | 19.03 | 0.26 | 0.98 | 0.77 | 0.85 | 0.78 |
| GFAP | 0.80(0.75-0.85) | 208.82 | 8.88 | 0.23 | 0.97 | 0.78 | 0.72 | 0.77 |
| NfL | 0.87(0.83-0.90) | 18.27 | 16.13 | 0.29 | 0.98 | 0.83 | 0.77 | 0.82 |

Abbreviations: PET, positron emission tomography; AUC, area under curve; OR, odds ratio; NPV, negative predictive value; PPV, positive predictive value; Sens., sensitivity; Spec., specificity; Accu., accuracy; CDR-GS, Clinical Dementia Rating Global Score. Thresholds were determined using the Youden index for biomarker‑only models.

**Table S3. Plasma biomarker performance for Aβ‑PET positivity, clinical diagnosis, all‑cause dementia, and 5‑ or 10‑year progression (biomarker + covariate models).**

|  | **AUC (95% CI)** | **OR** | **NPV** | **PPV** | **Sens.** | **Spec.** | **Accu.** |
| --- | --- | --- | --- | --- | --- | --- | --- |
| **Aβ-PET positive vs. Aβ-PET negative** | | | | | | | |
| p-tau181 | 0.79(0.72-0.84) | 7.15 | 0.70 | 0.75 | 0.73 | 0.72 | 0.73 |
| p-tau217 | 0.95(0.92-0.98) | 74.29 | 0.88 | 0.91 | 0.90 | 0.90 | 0.90 |
| GFAP | 0.84(0.78-0.88) | 11.43 | 0.75 | 0.79 | 0.78 | 0.77 | 0.77 |
| NfL | 0.72(0.64-0.78) | 5.20 | 0.64 | 0.75 | 0.63 | 0.75 | 0.69 |
| **Clinically unimpaired vs. AD dementia (Biomarker-only models)** | | | | | | | |
| p-tau181 | 0.84(0.82-0.85) | 11.20 | 0.56 | 0.90 | 0.78 | 0.76 | 0.78 |
| p-tau217 | 0.89(0.87-0.90) | 20.96 | 0.62 | 0.93 | 0.82 | 0.82 | 0.82 |
| GFAP | 0.85(0.83-0.87) | 11.99 | 0.57 | 0.90 | 0.80 | 0.75 | 0.79 |
| NfL | 0.85(0.83-0.87) | 12.03 | 0.56 | 0.90 | 0.78 | 0.77 | 0.78 |
| **Non-AD dementia vs. AD dementia** | | | | | | | |
| p-tau181 | 0.78(0.75-0.80) | 6.32 | 0.26 | 0.95 | 0.67 | 0.76 | 0.68 |
| p-tau217 | 0.80(0.77-0.82) | 6.80 | 0.28 | 0.94 | 0.72 | 0.73 | 0.72 |
| GFAP | 0.76(0.73-0.79) | 5.92 | 0.30 | 0.93 | 0.78 | 0.63 | 0.76 |
| NfL | 0.76(0.73-0.79) | 5.76 | 0.31 | 0.93 | 0.80 | 0.59 | 0.77 |
| **CDR-GS=0 vs. CDR-GS>=1** | | | | | | | |
| p-tau181 | 0.82(0.80-0.84) | 9.41 | 0.51 | 0.90 | 0.79 | 0.72 | 0.77 |
| p-tau217 | 0.86(0.85-0.88) | 14.28 | 0.57 | 0.91 | 0.82 | 0.75 | 0.81 |
| GFAP | 0.85(0.83-0.87) | 12.18 | 0.51 | 0.92 | 0.76 | 0.79 | 0.77 |
| NfL | 0.86(0.84-0.88) | 14.19 | 0.60 | 0.90 | 0.86 | 0.71 | 0.82 |
| **CDR-GS=0.5 vs. CDR-GS>=1** | | | | | | | |
| p-tau181 | 0.70(0.68-0.72) | 3.49 | 0.61 | 0.69 | 0.61 | 0.69 | 0.65 |
| p-tau217 | 0.73(0.71-0.74) | 4.20 | 0.64 | 0.70 | 0.67 | 0.68 | 0.67 |
| GFAP | 0.72(0.70-0.74) | 4.14 | 0.63 | 0.71 | 0.63 | 0.71 | 0.67 |
| NfL | 0.74(0.72-0.75) | 4.57 | 0.62 | 0.74 | 0.59 | 0.76 | 0.67 |
| **Progressors vs. non-progressors (5-year period)** | | | | | | | |
| p-tau181 | 0.80(0.77-0.83) | 7.94 | 0.46 | 0.90 | 0.68 | 0.79 | 0.71 |
| p-tau217 | 0.85(0.83-0.87) | 15.39 | 0.54 | 0.93 | 0.76 | 0.83 | 0.78 |
| GFAP | 0.82(0.80-0.85) | 9.87 | 0.56 | 0.88 | 0.82 | 0.68 | 0.79 |
| NfL | 0.83(0.81-0.86) | 10.96 | 0.52 | 0.91 | 0.75 | 0.78 | 0.76 |
| **Progressors vs. non-progressors (10-year period)** | | | | | | | |
| p-tau181 | 0.83(0.79-0.86) | 11.25 | 0.28 | 0.97 | 0.84 | 0.69 | 0.82 |
| p-tau217 | 0.89(0.86-0.92) | 20.97 | 0.24 | 0.98 | 0.75 | 0.88 | 0.76 |
| GFAP | 0.85(0.81-0.88) | 13.81 | 0.28 | 0.97 | 0.83 | 0.75 | 0.82 |
| NfL | 0.90(0.87-0.92) | 24.06 | 0.25 | 0.99 | 0.75 | 0.89 | 0.77 |

Abbreviations: PET, positron emission tomography; AUC, area under curve; OR, odds ratio; NPV, negative predictive value; PPV, positive predictive value; Sens., sensitivity; Spec., specificity; Accu., accuracy; CDR-GS, Clinical Dementia Rating Global Score. Thresholds were determined using the Youden index for biomarker‑only models. Covariates included age, sex, race, *APOE4* carriership, and education.

**Table S4.** **Characteristics of participants with longitudinal follow-up.**

| **Characteristics** | **Total** | **Normal  CDR-GS=0** | **MCI CDR-GS=0.5** | **Dementia CDR-GS>=1** |
| --- | --- | --- | --- | --- |
| Sample size | 2457 | 378 | 1096 | 983 |
| Years of follow-up | 3.0(3.8) | 6.2(6.7) | 3.2(4.0) | 2.1(2.0) |
| Age, y | 73.0(12.7) | 69.6(10.2) | 72.3(12.5) | 75.1(12.9) |
| Sex |  |  |  |  |
| Male | 1024(41.7) | 136(36.0) | 511(46.6) | 377(38.4) |
| Female | 1433(58.3) | 242(64.0) | 585(53.4) | 606(61.6) |
| Race |  |  |  |  |
| NHW | 2262(92.1) | 312(82.5) | 1025(93.5) | 925(94.1) |
| B/AA | 195(7.9) | 66(17.5) | 71(6.5) | 58(5.9) |
| Education, y |  |  |  |  |
| <13 | 953(38.8) | 88(23.3) | 319(29.1) | 546(55.5) |
| >=13 | 1504(61.2) | 290(76.7) | 777(70.9) | 437(44.5) |
| *APOE4* |  |  |  |  |
| Non carrier | 1248(50.8) | 245(64.8) | 574(53.4) | 429(43.6) |
| Carrier | 1177(47.9) | 128(38.9) | 502(45.8) | 547(55.6) |
| Missing | 32(1.3) | 5(1.3) | 20(1.8) | 7(0.7) |
| Clinical diagnosis |  |  |  |  |
| Normal | 513(20.9) | 356(94.2) | 156(14.2) | 1(0.1) |
| MCI | 534(21.7) | 18(4.8) | 494(45.1) | 22(2.2) |
| AD | 1232(50.1) | 1(0.3) | 372(33.9) | 859(87.4) |
| Non-AD dementia | 174(7.1) | 3(0.8) | 72(6.6) | 99(10.1) |
| Missing | 4(0.2) | 0(0) | 2(0.2) | 2(0.2) |
| p-tau181, pg/ml | 45.4(30.7) | 33.3(19.6) | 43.0(29.3) | 53.0(30.5) |
| p-tau217, pg/ml | 0.76(0.97) | 0.37(0.35) | 0.65(0.83) | 1.14(1.09) |
| GFAP, pg/ml | 274.5(238.1) | 179.4(141.0) | 246.0(218.8) | 351.5(259.5) |
| NfL, pg/ml | 25.3(19.2) | 17.4(12.3) | 22.6(15.7) | 32.4(20.4) |

Abbreviations: CDR-GS, Clinical Dementia Rating Global Score; NHW, Non-Hispanic White; B/AA, Black/African American; AD, Alzheimer’s Disease; MCI, Mild Cognitive Impairment. The median and interquartile range (IQR) are reported for continuous variables, while frequencies are shown for categorical variables.

**Table S5.** **Characteristics of participants with 5- or 10-year progressing status.**

| **Characteristics** | **5-year period** | | | | **10-year period** | | | |
| --- | --- | --- | --- | --- | --- | --- | --- | --- |
|  | **Total** | **Non-progressor** | **Progressor** | **p** | **Total** | **Non-progressor** | **Progressor** | **p** |
| Sample size | 1368 | 345 | 1023 |  | 1250 | 106 | 1144 |  |
| Years of follow-up | 4.0(4.6) | 7.9(4.5) | 3.0(2.3) | 1.28E-123 | 3.4(4.1) | 13.0(4.8) | 3.1(3.3) | 3.28E-62 |
| Age, y | 72.9 (12.8) | 68.2(10.7) | 74.4(12.1) | 3.36E-22 | 73.8(12.5) | 66.2(10.2) | 74.3(11.7) | 1.91E-14 |
| Sex |  |  |  | 0.03 |  |  |  |  |
| Male | 547 (40.0) | 155 (44.9) | 392 (38.3) |  | 493(39.4) | 47(44.3) | 446(39.0 |  |
| Female | 821 (60) | 190 (55.1) | 631 (61.7) |  | 757(60.6) | 59(55.7) | 698(61.0) |  |
| Race |  |  |  | 0.001 |  |  |  | 0.018 |
| NHW | 1273(93.1) | 308(89.3) | 965(94.3) |  | 1174(93.9) | 94(88.7) | 1080(94.4) |  |
| B/AA | 95(6.9) | 37(10.7) | 58(5.7) |  | 76(6.1) | 12(11.3) | 64(5.6) |  |
| Education, y |  |  |  | 2.22E-16 |  |  |  | 4.04E-09 |
| <13 | 542(39.6) | 72(20.9) | 470(45.9) |  | 526(42.1) | 16(15.1) | 510(44.6) |  |
| >=13 | 826(60.4) | 273(79.1) | 553(54.1) |  | 724(57.9) | 90(84.9) | 634(55.4) |  |
| *APOE4* |  |  |  | 4.00E-13 |  |  |  | 8.36E-05 |
| Non carrier | 693(50.7) | 234(68.8) | 459(44.9) |  | 582(46.6) | 69(65.1) | 513(44.8) |  |
| Carrier | 667(48.8) | 111(32.2) | 556(54.4) |  | 660(52.8) | 37(34.9) | 623(54.5) |  |
| Missing | 8(0.6) | 0(0) | 8(0.8) |  | 8(0.64) | 0 | 8(0.7) |  |
| Clinical diagnosis |  |  |  | 0 |  |  |  | 0 |
| Normal | 250(18.3) | 206(59.7) | 44(4.3) |  | 151(12.1) | 71(67.0) | 80(7.0) |  |
| MCI | 240(17.5) | 107(31.0) | 133(13.0) |  | 203(16.2) | 29(27.4) | 174(15.2) |  |
| AD | 772(56.4) | 21(6.1) | 751(73.4) |  | 791(63.3) | 4(3.8) | 787(68.8) |  |
| Non-AD dementia | 106(7.7) | 11(3.2) | 95(9.3) |  | 104(8.3) | 2(1.9) | 102(8.9) |  |
| Missing | 0(0) | 0(0) | 0(0) |  | 1(0.1) | 0(0) | 1(0.1) |  |
| CDR-GS |  |  |  | 0 |  |  |  | 0 |
| 0 | 200(14.6) | 147(42.6) | 53(5.2) |  | 135(10.8) | 50(47.2) | 85(7.4) |  |
| 0.5 | 609(44.5) | 174(50.4) | 435(42.5) |  | 543(43.4) | 53(50.0) | 490(42.8) |  |
| 1 | 472(34.5) | 20(5.8) | 452(44.2) |  | 487(39.0) | 3(2.83) | 484(42.3) |  |
| 2 | 87(6.4) | 4(1.2) | 83(8.1) |  | 85(6.8) | 0(0) | 85(7.4) |  |
| p-tau181, pg/ml | 46.4(29.8) | 34.8(18.3) | 51.6(30.0) | 9.76E-37 | 49.5(29.8) | 34.3(17.6) | 51.2(30.0) | 2.44E-14 |
| p-tau217, pg/ml | 0.83(1.00) | 0.36(0.30) | 1.07(0.99) | 3.88E-71 | 0.93(1.02) | 0.29(0.23) | 1.03(0.99) | 2.09E-35 |
| GFAP, pg/ml | 284.9(235.8) | 174.7(145.5) | 326.1(242.6) | 5.38E-56 | 304.4(238.0) | 155.6(123.3) | 318.6(237.0) | 4.21E-25 |
| NfL, pg/ml | 26.3(19.1) | 16.3(13.1) | 29.4(20.1) | 1.23E-56 | 27.4(19.7) | 13.7(8.2) | 28.7(19.7) | 2.20E-36 |

Abbreviations: CDR-GS, Clinical Dementia Rating Global Score; NHW, Non-Hispanic White; B/AA, Black/African American; AD, Alzheimer’s Disease; MCI, Mild Cognitive Impairment. The median and interquartile range (IQR) are reported for continuous variables, while frequencies are shown for categorical variables.
